# New primer sets to detect recent human adenovirus F41 variants in wastewater: Is it linked to the new acute hepatitis?

**DOI:** 10.1101/2022.09.16.22280038

**Authors:** Mariel Perez-Zabaleta, Cecilia Williams, Zeynep Cetecioglu

**Affiliations:** Department of Industrial Biotechnology, KTH Royal Institute of Technology, AlbaNova University Center, SE-10691, Stockholm, Sweden; Department of Protein Science, KTH Royal Institute of Technology, Science for Life Laboratory, Solna, Sweden

**Keywords:** Human adenovirus F41 (HAdV-F41), human adenovirus F40 (HAdV-F40), wastewater-based epidemiology (WBE), acute hepatitis, pepper mild mottle virus (PMMoV)

## Abstract

Human adenovirus type F-41 has been pursued as one of the potential reasons for the new acute hepatitis cases of unknown cause in young children. Tracking the spread of this virus in the population using wastewater-based epidemiology tools can help clinical investigations to determine its relation to this new hepatitis outbreak.

In this study, methods to detect human adenovirus type F (40 and 41) and specifically type F41 were designed and implemented to quantify the amount of these pathogens in wastewater samples from Stockholm, Sweden. An assay based on reverse transcriptase quantitative polymerase chain reaction using TaqMan technology and primers targeting the three main capsid genes of adenoviruses: hexon, penton and fiber, was designed. The hexon primers were specific to adenovirus F41, while fiber primers could quantify both adenoviruses, F40 and F41. Wastewater samples from Stockholm were used to validate the designed assay and, in addition, pepper mild mottle virus (PMMoV) levels were quantified to study the data normalization.

Our results can help link the occurrence of the virus variant with new cases of acute hepatitis and contribute to a better understanding of the possible causes. It can also provide valuable information that can be used in future investigations on the monitoring of human adenovirus type F in wastewater.

## 1 Introduction

Human adenoviruses are nonenveloped DNA viruses belonging to the family Adenoviridae. These viruses are classified into seven species (A to G) and at least 104 genotypes (HAdV Working Group, 2022). Adenoviruses can cause respiratory illness, gastroenteritis, conjunctivitis, or cystitis in young children (Ikner and Gerba, 2017). There is no specific treatment for adenovirus infections. Adenovirus type F (40 and 41) usually causes acute gastroenteritis in children. However, cases of hepatitis have been reported in immunocompromised children associated with adenovirus type 41 (HAdV-F41) infection (Ikner and Gerba, 2017).

A sudden increase in acute hepatitis cases of unknown cause was reported to the World Health Organization (WHO) in 2022. Between April 5 and July 8, 2022, 1 010 probable cases and 22 deaths were reported by 35 countries, including five pediatric patients with severe hepatitis of unknown origin identified at a children’s hospital in Alabama, U.S., in early October 2021 (WHO, 2022). Of the 1 010 probable cases, 5% of the children required a liver transplant, and almost half of the cases (48%) were reported from European countries with the UK accounting for the highest number. The children were under 16 years old, presented with liver inflammation (including some with liver failure), and tested negative for hepatitis A to E (WHO, 2022). The cases were not associated with travel, and most children tested positive for adenovirus, in which type 41 was predominant. Of 251 cases tested for adenovirus in the UK, 68% gave positive results against this pathogen (UK Health Security Agency, 2022). In Alabama, HAdV-F41 was detected in all five infected patients (Baker et al., 2021). A recent investigation has pointed to the synergism between HAdV-F41 and SARS-CoV-2 as responsible to activate higher levels of inflammation, which eventually can cause acute hepatitis in children (Luan et al., 2022).

Wastewater-based epidemiology (WBE) is a useful tool to detect the occurrence of viruses in a population, as viruses do not possess the ability to grow outside their host and their concentration in wastewater can be representative of the infection rate (Xagoraraki and O’brien, 2020). The wastewater-based epidemiology concept was initially perceived in 2001 (Daughton C.G. and Jones-Lepp T.L., 2001) and applied to grasp the circulation of viruses in previous outbreaks such as poliovirus in the Netherlands or hepatitis A and norovirus in Sweden (Hellmér et al., 2014). Simultaneous detection of different adenoviruses genotypes was previously investigated to determine the removal efficiency of “generic” adenoviruses through the wastewater treatment process (Carducci et al., 2008; Hata et al., 2012; Quintão et al., 2021). Few studies were able to specifically identify HAdV-F41 (Heim et al., 2003; Kuo et al., 2009; Tiemessen and Nel’, 1996), and previously designed primers to detect HAdV-F41 can be less specific for emergent variants. According to Götting and colleagues (2022), HAdV-F41 has evolved significantly over the past decade, which possibly has resulted in altered virulence of this pathogen. The emergence of a novel strain of HAdV-F41 could be suspected, which may be causing hepatitis. Unfortunately, recovery of high-quality HAdV-F41 genomes from hepatitis-affected children has failed due to low virus loads (Kajon and St George, 2022).

In relation to the genetic diversity of adenoviruses, sequencing from patient samples revealed that the genes of hexon, fiber and penton include hypervariable regions that can be used for genotyping (Risso-Ballester et al., 2016). Fiber, penton, and hexon as the main capsid proteins of adenoviruses are responsible for binding to the host cell. (Larson et al., 2015).

In this study, the gene sequences of these three major capsid genes, hexon, penton and fiber, were used to design the primers and probes for the quantification of adenovirus type F by RT-qPCR. The difference between the specificity of the primers for different variants was tested. Wastewater samples from Stockholm were used to validate the designed methods. Contents of human adenovirus F40 (HAd-V-F40) and HAdV-F41 were detected in the wastewater samples. In addition, pepper mild mottle virus (PMMoV) was also determined in the samples and studied for data normalization.

## 2 Materials and Methods

### 2.1 Design of primers to detect human adenovirus type F and in silico validation

Primers and probes targeting the main capsid genes of adenovirus: hexon, penton and fiber, were designed using recently published genome sequences of HAdV-F41 variants current in 2021 and 2022: MW567962.1 (March 2021), ON532826.1 (May 2022) and OP174922.1 (August 2022). The primers and probes are listed in Table 1.

**Table 1.**
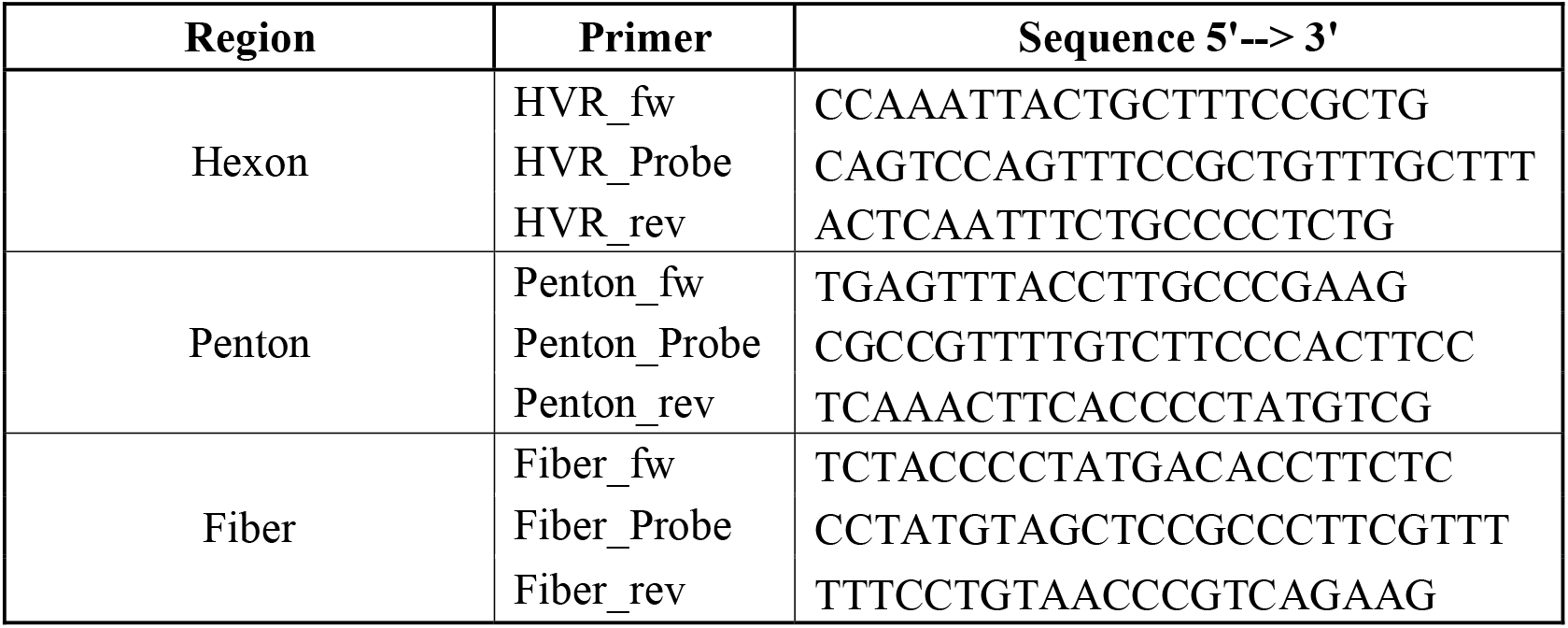
List of primer sequences used to quantify Human Adenovirus type F.

The primers were analyzed in silico using Basic Local Alignment Tool (BLAST) from the National Center for Biotechnology Information (NCBI) and the sequences of the three primer sets were confirmed to match HAdV-F41. Then, the primer sequences were aligned against twenty HAdV-F41 species and two HAdV-F40 species from NCBI GenBank. The alignments were performed using DNASTAR Bioinformatics Software. The twenty HAdV-F41 genomes were selected according to a recent phylogenetic study, which indicated that HAdV-F41 evolved with three lineages (Götting et al., 2022). This recent investigation analyzed 65 genomes from which Lineage 1 included HAdV-F41 prototype strain Tak from 1973 (GenBank DQ315364) and six isolates from 2007 to 2017. Lineage 2 included 53 isolates of the 65 genomes studied and strains from 2000 to 2022. Lineage 3 included 6 isolates and this group was closer to the prototype strain HAdV-F40 Dugan, isolated in 1979. The selected HAdV-F41 genomes were from lineage 1 and 2. In lineage 1, we included the Tak strain, while lineage 2 included HAdV-F41 genomes used in this investigation (MW567962.1, ON532826.1, and OP174922.1) to design the primers/probes.

### 2.2 In vitro validation of primers to detect human adenovirus type F

The hexon, penton, and fiber primers were evaluated against thirteen genotypes of human adenovirus for species A, B, C, D and E (Table 3). In addition, three positive controls of adenovirus type F were tested. Two variants of HAdV-F41, one prototype strain Tak (GenBank: DQ315364.2) and one strain isolated from a Swedish patient (GenBank: KX868523.2) (Lá szló Kajá et al., 2018), and a positive control for HAd-V-F40 (the prototype strain HoviX, GenBank: KU162869.1) were also tested. The adenovirus species were provided by the Norrlands Universitetssjukhus (Klinisk Mikrobiologi, Umeå, Sweden). Adenovirus controls were heat inactivated at 56°C for 30 min and then, 200 μL of each culture was used for DNA extraction using Maxwell RSC Enviro TNA Promega Kit and Maxwell RSC Instrument (Promega Biotech AB, Sweden) following the manufacturer’s instructions.

### 2.3 Reverse transcriptase quantitative polymerase chain reaction (RT-qPCR) method using the designed primer sets

RT-qPCR assays were performed using 5 μL of TaqMan Reliance One-Step Multiplex Supermix (BioRad, Cat # 12010221). 1 μL of 20 mg/ml Bovine Serum Albumin (BSA) (Thermo Scientific, Cat # B14) was added to reduce PCR inhibitors and enhance efficacy, for a final reaction volume of 20 μL. For each reaction, 5 μL of DNA template was used. Nuclease-free water and DNA extracted from tap water were included as negative controls for all qPCR reactions. Thermal cycling (50 °C 10 min, 95 °C 10 s, followed by 45 cycles of 95 °C 3 s, 60 °C for 30 s) was performed on an Applied Biosystems QuantStudio 3 RT-qPCR machine (ThermoFisher Scientific). Reactions were considered positive if the cycle threshold (Ct) was below 42 cycles. Amplifications were carried out on the FAM channel.

PMMoV was also quantified in the samples to study data normalization. The PMMoV primers used were forward: 5’-GAGTGGTTTGACCTTAACGTTTGA-3’ reverse: 5’-TTGTCGGTTGCAATGCAAGT-3’ and probe 5’-CCTACCGAAGCAAATG-3’ (Zhang et al., 2006).

Standard curves were created using constructed plasmids containing appropriate regions for hexon, penton, fiber, and PMMoV (IDT, Custom MiniGene 25-500 bp).

### 2.4 Validation of primer sets in wastewater samples

#### 2.4.1 Wastewater samples

The validation of the primer sets was performed in wastewater samples from Stockholm, which is the capital and largest city of Sweden and accounts for a population of approx. 2.4 million people in the metropolitan area. The wastewater samples were collected from the three main municipal wastewater treatment plants (WWTP): Stockholm Vatten och Avfall Bromma, Stockholm Vatten och Avfall Henriksdal, and Käppala WWTP. Bromma WWTP treats wastewater from approximately 377 500 inhabitants and samples were obtained from three inlets (corresponding to three regions: Hässelby, Riksby, and Järva). The second analyzed WWTP, Henriksdal, treats wastewater from 862 100 inhabitants and has two inlets, Sickla and Henriksdal. Käppala WWTP covers a region of 700 000 inhabitants and has one inlet.

Approximately 500 ml of wastewater samples from each inlet were taken over a period of 24 hours from Monday to Tuesday, using a flow-proportional composite sampler. Wastewater samples were transported to the laboratory on ice and kept at 4°C until further analyses. After receiving the samples, two representative samples for the whole Stockholm region were attained by mixing the different inlets according to the percentage of flow rates of each region (Table 2). The representative samples from Stockholm were further concentrated and analyzed.

**Table 2.** Flow rates and inhabitants of the WWTPs from Stockholm. Average flow rates were calculated using historical data of flow rates from April 2020 to June 2022.

| WWTP | Inlet | Average<br>flow rate<br>(m<sup>3</sup>/s) | Inhabitants | Flow<br>rate (%) |
| --- | --- | --- | --- | --- |
| **Henriksdal** | Henriksdal | 1.52 | 371800 | 23% |
|  | Sickla | 1.84 | 490200 | 28% |
| **Bromma** | Hässelby | 0.44 | 125800 | 7% |
|  | Järva | 0.78 | 197200 | 12% |
|  | Riksby | 0.24 | 37500 | 4% |
| **Käppala** | Käppala | 1.76 | 700000 | 27% |

**Table 3.** Specificity test of hexon, penton and fiber primer-sets toward various human adenovirus species.

| Adenovirus Control | Species | Classification | RT-qPCR results |  |  |
| --- | --- | --- | --- | --- | --- |
|  |  |  | Hexon | Penton | Fiber |
| Adenovirus 31 | A | A31 | Negative | Negative | Negative |
| Adenovirus 3 | B1 | B3 | Negative | Negative | Negative |
| Adenovirus 7 | B1 | B7 | Negative | Negative | Negative |
| Adenovirus 11 | B2 | B11 | Negative | Negative | Negative |
| Adenovirus 14 | B2 | B14 | Negative | Negative | Negative |
| Adenovirus 34 | B2 | B34 | Negative | Negative | Negative |
| Adenovirus 1 | C | C1 | Negative | Negative | Negative |
| Adenovirus 2 | C | C2 | Negative | Negative | Negative |
| Adenovirus 5 | C | C5 | Negative | Negative | Negative |
| Adenovirus 9 | D | D9 | Negative | Negative | Negative |
| Adenovirus 13 | D | D13 | Negative | Negative | Negative |
| Adenovirus 20 | D | D20 | Negative | Negative | Negative |
| Adenovirus 4 | E | E4 | Negative | Negative | Negative |

#### 2.4.2 Wastewater concentration and DNA extraction

Wastewater viral concentration and total nucleic acid (TNA) extraction were usually performed the same or the next day after receiving the samples. Concentration and TNA extraction were performed using Maxwell RSC Enviro TNA Promega Kit and Maxwell RSC Instrument (Promega Biotech AB, Sweden). High concentrations and yields were reported when this kit has been used in wastewater samples (Perez-Zabaleta et al., 2022), (Isaksson et al., 2022). Briefly, 40 ml of wastewater was treated with a protease solution and centrifugated at 3000 g for 10 min to remove proteins and solids present in the wastewater. Then, the supernatant was filtrated, eluted to 500 L using a column-based system, and then loaded into a cartridge provided by the kit. Maxwell RSC Pure Food GMO program was selected in the instrument software for automatic TNA extraction and eluted in 80 μL nuclease-free water. Two independent replicates were analyzed for each sample and tap water was used as a negative control for viral concentration and DNA extraction steps.

#### 2.4.3 Sanger sequencing of PCR amplicons from wastewater samples

The PCR amplicons from the wastewater samples obtained after RT-qPCR were purified using GeneJET PCR Purification Kit (ThermoFisher Scientific, Cat # K0701) following the manufacturer’s instructions. Two pure PCR products per sample were sent for sequencing, one PCR product was sequenced in the forward direction and the other one in the reverse direction. Sanger sequencing was performed by Eurofins using Mix2Seq Kit.

To determine the HAdV genotypes in the wastewater samples, hexon, penton and fiber sequences obtained by Sanger sequencing were compared with sequences in the NCBI databases by using BLAST.

## 3 Results

### 3.1 Primers and probes do not amplify a panel of related human adenoviruses

The primers and probes were designed to be specific to adenovirus type F. To test their specificity, different adenovirus species (A, B, C, D and E) were used as negative controls. In total, thirteen adenovirus genotypes were analyzed (Table 3). The three sets of primers (hexon, penton and fiber) gave negative results for species A to E. Thus, they should not detect these species.

### 3.2 Primers and probes amplify adenovirus type F

Next, the primers and probes were tested for adenovirus type F. One positive control for HAdV-F40 (strain HoviX) and two variants of HAdV-F41 (strain Tak and strain isolated from a Swedish patient) were analyzed and used to validate the primer sets in vitro.

Two primer sets (fiber and penton) detected the HAdV-F40 (strain HoviX). The amount detected by the fiber primer-set was almost 8000 times higher than the detected by the penton primers. Hexon primers gave negative results, showing no specificity towards HAdV-F40 (Figure 1a). The amount of gene copy numbers of HAdV-F40 (strain HoviX) per μL was significantly different (P-value = 0.0007) among the three primer sets. For adenovirus F41 (strain Tak), the penton or fiber primer sets detected similar amounts and were not significantly different (P-value = 0.30, Figure 1b). The primers targeting the hexon region could not detect this strain either. Alignments of the genome of HAdV-F41 strain Tak and hexon primers showed that forward and reverse primers could bind the hexon region but the designed probe (HVR_probe) had in total seven mismatches in the nucleotides, in which four of them were consecutive. However, the HAdV-F41 variant previously isolated from a Swedish patient was detected by all three primer sets, which quantified similar amounts of the virus, with no significant difference (P-value = 0.35, Figure 1c).

**Figure 1.**
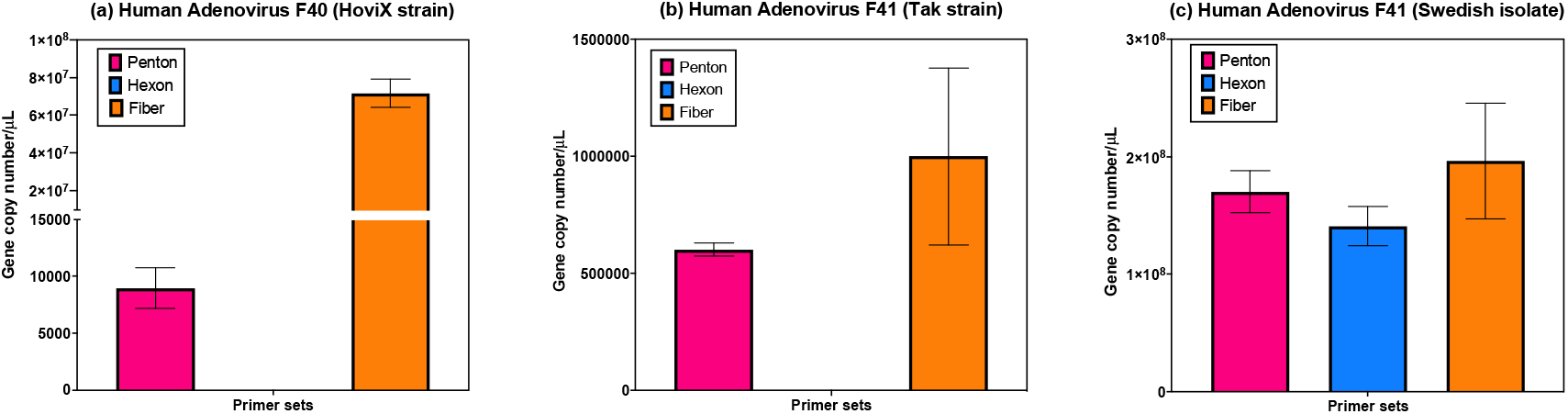
Analysis of positive controls for human adenovirus F40 and F41 using primers-sets for hexon, penton and fiber regions. Detection and gene copy numbers (per μL) of HAdV-F40 and F41 were determined by qPCR analysis of the hexon, penton or fiber region in three controls (a) Prototype HAdV-F40 HoviX strain (b) Prototype HAdV-F41 Tak strain (c) and an HAdV-F41 strain isolated from a patient in Sweden.

The validation of primers in silico and in vitro showed that hexon primers are selective for recent HAdV-41 strains evolved in lineage 2. Fiber and penton primers can be used to detect both HAdV-40 and HAdV-41 variants and detect the HAdV-41 Swedish variant at the same sensitivity as hexon primers (but are not selective for this variant). Comparing penton and fiber primers, penton is less sensitive to HAdV-40 than fiber primers, however, both primer sets can detect similar amounts of both HAdV-41 strains (Figure 1b and Figure 1c).

### 3.3 Validation of primer sets using wastewater samples

#### 3.3.1 Quantification of Human Adenovirus type F

Next, the primer sets were tested for the detection of human adenovirus F in Stockholm wastewater. Wastewater samples from six inlets (three WWTPs) were mixed proportionally to the flow rates to obtain a representative Stockholm sample. Two biological replicates were analyzed by RT-qPCR with the three primer sets (hexon, penton and fiber) as well as with PMMoV primer set, and quantified. The results obtained from the wastewater samples showed that primers targeting the hexon region detected lower amounts of adenovirus compared to penton or fiber primers (Figure 2). These results are in line with the outcomes obtained in the specificity study (section 3.2) since hexon primers were more specific to HAdV-41 lineage 2 and did not detect HAdV-F40 or HAdV-F41 Tak strain. Fiber primers detected higher levels of adenovirus compared to penton primers in all the samples (Figure 2b). In all the analyzed samples, the fiber and penton assays detected the same trends of increasing or decreasing amounts of virus, whereas the hexon primers detect a different trend (Figure 2b). The fiber and penton assays also both detected about 100 times more viral DNA copies (around 10^17^) than the hexon primer (10^15^). This suggests that the wastewater samples contain a mixture of HAdV-F strains (detected by fiber and penton) and strains similar to the HAdV-F41 Swedish isolate (detected selectively by the hexon assay) make up 1% or less or the circulating HAdV-F40 and 41 strains. However, this estimation is uncertain as it cannot account for potential variations in the sequences of the primer and probe sites among developing variants which would impact amplification efficacy or detection.

**Figure 2.**
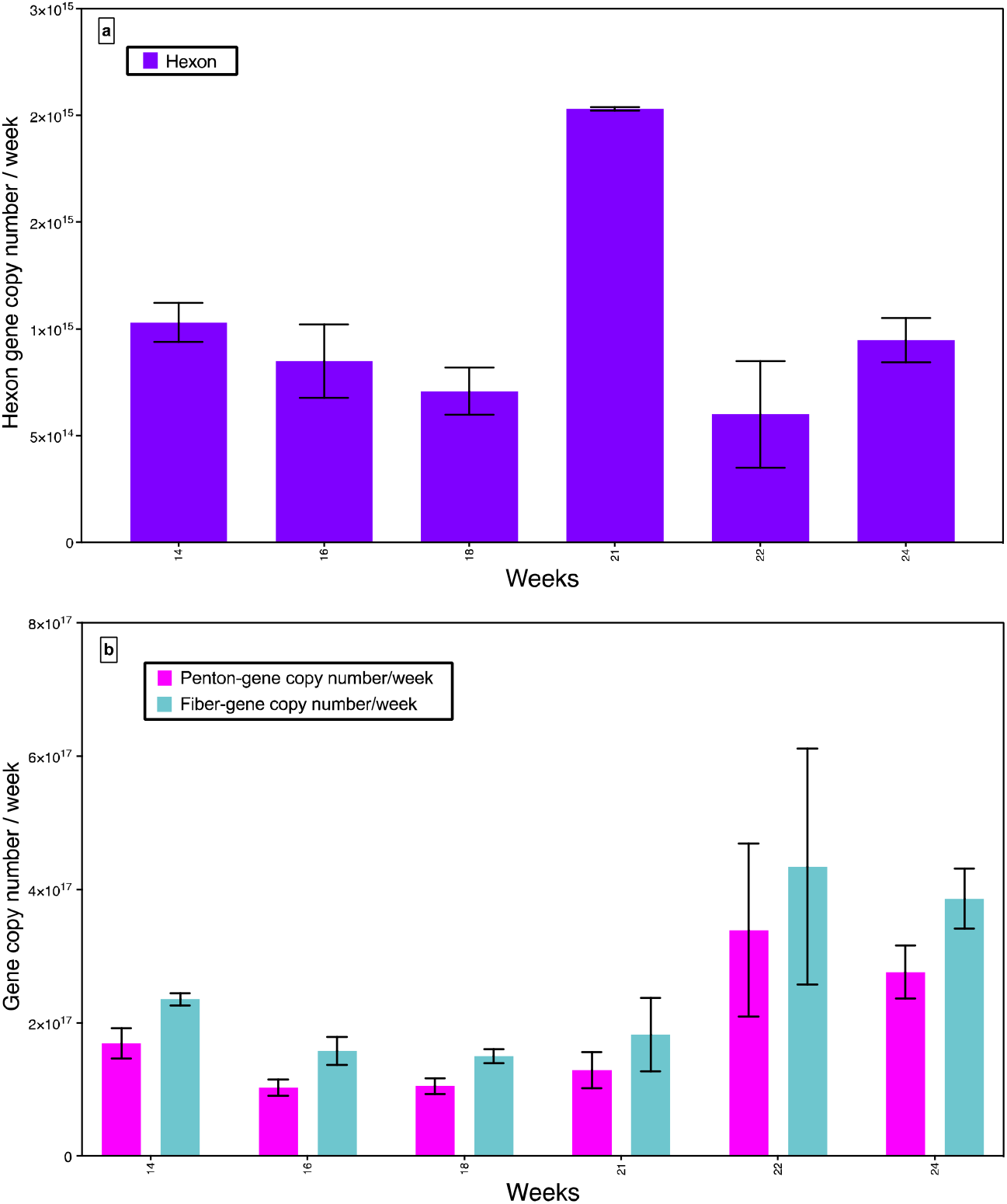
Quantification of HAdV-F40 and F41 using primers-sets for hexon, penton and fiber regions in wastewater samples. Gene copy numbers of HAdV-F41 and F40 per μL were determined by targeting (a) hexon region (purple), (b) penton (pink) and fiber (green) regions. Two biological replicates were analyzed for each sampling point.

#### 3.3.2 Sanger sequencing of PCR products from wastewater samples

To determine the specific HAdV genotypes detected by the assays in the wastewater samples, the amplified hexon, penton and fiber PCR products were analyzed by Sanger sequencing. The obtained sequences were compared in the NCBI databases by BLAST. The DNA sequences from the PCR products analyzed by hexon primers were specific for HAdV-F41, which confirms that the hexon method only detects adenovirus F41. In the case of penton and fiber, the DNA sequences were not specific to HAdV-F41 and a double detection of HAdV-F40 and HAdV-F41 can be suspected.

#### 3.3.3 Normalization of human adenovirus type F detection to populations

The size of a population in a specific area is a variable factor difficult to predict. The PMMoV is an RNA virus that is abundant in human faeces and has been reported to be a good population indicator in wastewater samples (Kitajima et al., 2018; Zhang et al., 2006). Furthermore, in our previous investigations of SARS-CoV-2 in wastewater, PMMoV was found to be an effective normalization factor to correct for fluctuations in the wastewater content (Perez-Zabaleta et al., 2022). However, the normalization of adenovirus to PMMoV has not yet been explored.

Comparing our data for HAdV-F with and without normalization to PMMoV, it can be observed that the trends of increasing or decreasing levels are different (Figure 3 compared to Figure 2). The PMMoV-normalized data show more constant values among the analyzed weeks for the three assays (hexon, penton and fiber primer sets, Figure 3). While non-normalized data indicated a sudden peak of HAdV-F41 (Stockholm variant) in week 21, the normalized data show a higher level during spring (week 14-21) that decreases toward the end of May. Similarly, the overall HAdV-F40 and 41 variants appear to decrease during May. As adenoviruses are more common in the Scandinavian regions during winter and early spring, this may suggest that PMMoV normalization can help to decrease fluctuation in the measurement due to population size. However, to corroborate this, the trends should be correlated to the actual number of clinical cases. Unfortunately, a such comparison was not possible since the number of infected persons by adenovirus type F per week in Stockholm is unknown.

**Figure 3.**
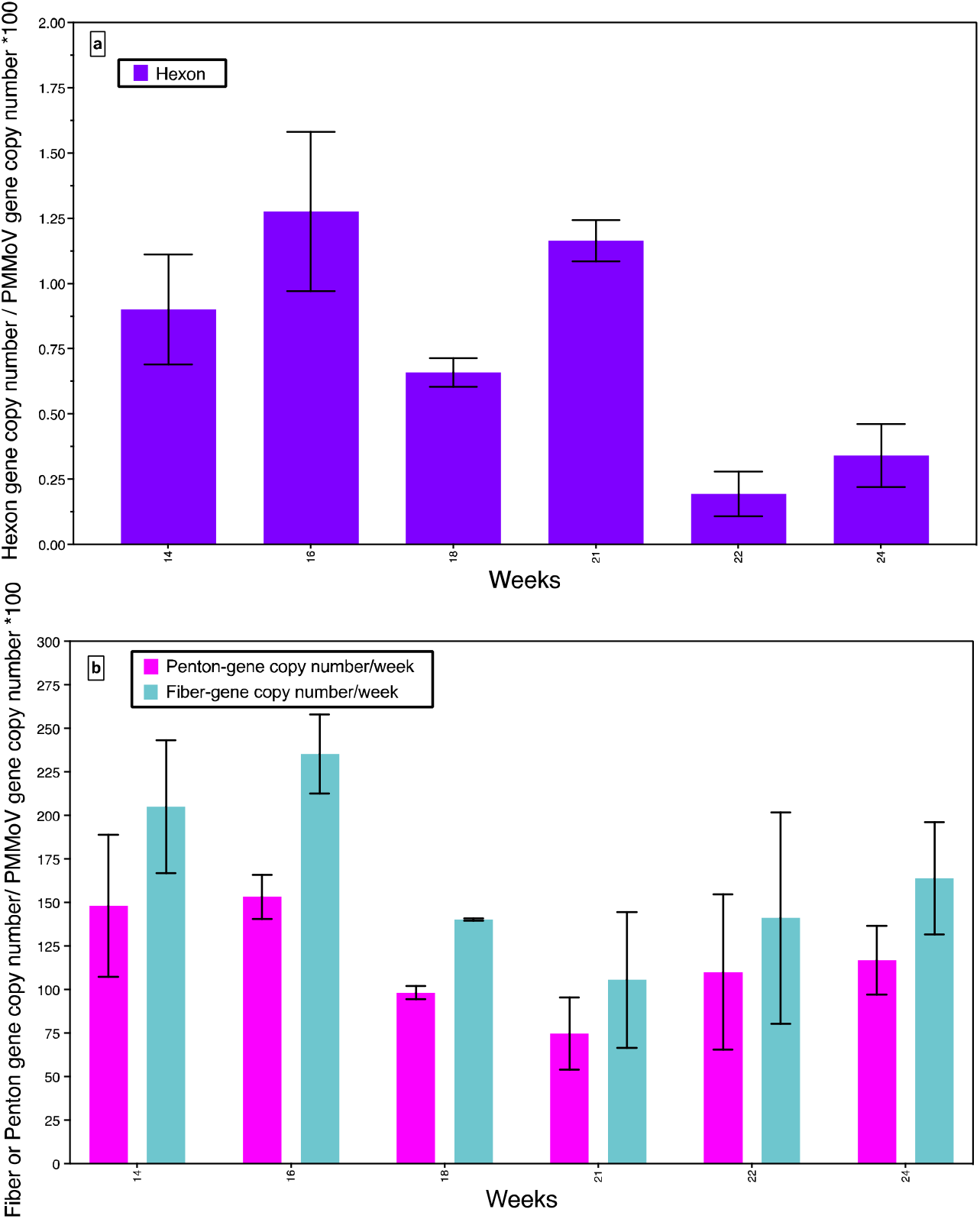
Normalization of HAdV-F40 and F41 in wastewater samples with respect to PMMoV. Samples were analyzed using primers sets for hexon, penton and fiber regions. PMMoV content was determined and used to normalize the data. The amount of adenovirus is presented as (a) hexon (purple) gene copy numbers per PMMoV gene copy number *100, (b) penton (pink) or fiber (green) gene copy numbers per PMMoV gene copy number *100. Two biological replicates were analyzed for each sampling point.

## Conclusion

- Hexon primer sets are more specific to HAdV-41, especially from lineage 2.
- Fiber primer sets can detect both HAdV-40 and HAdV-41.
- PMMoV may be appropriate for the normalization of HAdV measurements in wastewater but further studies are needed.
- Routine sequencing of PCR products from wastewater could provide detailed information on HAdV variant spread.

## Data Availability

All data produced in the present work are contained in the manuscript

## Author approval

All authors interpreted the data, revised the manuscript, and approved the final version of the manuscript.

## Acknowledgements

The authors would like to thank the Science for Life Laboratories Environmental Virus Profile Platform, Swedish Environmental Epidemiology Center (SEEC), the KaDppala Association, and Stockholm Vatten och Avfall. Additionally, the authors acknowledge SEEC collaborators for project management, and Annika Alland from Norrlands Universitetssjukhus, Klinisk Mikrobiologi, Umeå, Sweden for providing positive controls of different adenovirus genotypes and scientific discussions.

